# Automated AI-Based Ventricular Subcompartment Segmentation and Volumetry in Idiopathic Normal Pressure Hydrocephalus

**DOI:** 10.64898/2026.06.14.26355627

**Authors:** Matthias Anthony Mutke, Stephanie Ariane Griot, Jakob Wasserthal, Ashraya Kumar Indrakanti, Nathan Vishwanathan, Mustafa Ahmed Mahmutoglu, Tugba Akinci D’Antonoli, Michael Bach, Marios Psychogios, Johanna Maria Lieb

## Abstract

**Purpose:** In idiopathic normal pressure hydrocephalus (iNPH), longitudinal monitoring of ventricular size is important for diagnosis and treatment follow-up. This study aimed to validate a fully automated AI model for CT ventricular volumetry with subcompartments and to compare AI-derived volume changes with routine radiology assessments.

**Methods:** This retrospective, single-center study included 88 patients with iNPH and 456 non–contrast-enhanced head CT examinations. The model was trained on 38 manually labeled CT scans with 12 ventricular subcompartments. Outcomes included segmentation accuracy, correspondence between AI-derived longitudinal ventricular volume changes and radiology report categories (decreased, unchanged, increased), radiologist detection thresholds for ventricular change, and paired pre- and postoperative volume changes in 22 patients with ventriculoperitoneal shunt.

**Results:** Mean segmentation accuracy was high (Dice, 0.83). 91% of 100 segmentations were rated as excellent by an expert neuroradiologist. AI-derived ventricular volume changes corresponded well to radiology report categories (median total ventricular volume changes of −17% in cases reported as decreased, 0% in unchanged cases, and +22% in increased cases (all p < 0.001)). Radiologists reported ventricular volume change in 50% of cases at an AI-measured relative volume change of ±6%, and in 90% of cases at +21% for enlargement and −18% for decrease. After shunt placement, ventricular volume decreased by −8% (median), with the largest relative reductions observed in the right temporal and occipital horns.

**Conclusions:** Automated AI-based ventricular segmentation on CT enables accurate and reproducible assessment of ventricular volume changes in iNPH and complements routine radiological evaluation for longitudinal and postoperative monitoring.

## Introduction

Determining ventricular size and its longitudinal change in idiopathic normal pressure hydrocephalus (iNPH) is important for evaluating ventriculomegaly, establishing diagnostic criteria, and assessing the effectiveness of shunt treatment[1].

Traditionally, ventricular size and change has been estimated through subjective visual inspection or by using biplane measurements such as the Evans index (EI) on imaging[2]. However, visual assessment is inherently limited, and the accuracy of the EI is influenced by factors such as image selection, head positioning, and interindividual anatomical variation[3]. Also, qualitative radiological assessment does not reliably predict shunt response[4].

These limitations have driven the development of quantitative volumetric methods that provide more objective and reproducible assessments of ventricular size[5]. Recent advances in deep learning–based automated segmentation have the potential to deliver higher accuracy, consistency, and applicability, even in postoperative conditions where imaging artifacts are common[6]. Further segmentation of ventricular subcompartments could enable more precise localized evaluation of the anatomical distribution of potential changes. Importantly, there is a growing need for open-source models evaluated in real-world clinical settings to facilitate translation into clinical practice[7].

The aim of this study was therefore to develop and clinically evaluate an open-source AI model for longitudinal ventricular volume measurement including pre- and postoperative CT scans in a clinical cohort of iNPH patients with accurate subcompartment segmentation.

## Materials and methods

### Study Design and model availability

This retrospective diagnostic imaging study evaluated the accuracy and clinical usefulness of a fully automated artificial intelligence (AI)–based ventricular segmentation and volume measurement tool in patients with idiopathic normal pressure hydrocephalus (iNPH). The model is a customized nnU-Net trained from scratch on manually labeled CT scans and integrated into the TotalSegmentator framework. The study adheres to the CLAIM guidelines[8].

The study was conducted at a single tertiary care hospital in central Europe using fully anonymized data. Informed consent was waived by the local ethics committee. The model is available as open-source software via the TotalSegmentator project (https://github.com/wasserth/TotalSegmentator/; https://totalsegmentator.com/).

### Study Population

We retrospectively reviewed the medical records and imaging data of patients with documented clinical diagnosis of probable or possible iNPH between January 14, 2011, and November 15, 2022. Imaging datasets were identified using an in-house developed open-source PACS search engine[9] utilizing relevant diagnostic codes and keywords.

The dataset was split into a training set (n=38 CT scans) and a test set (n=456 CT scans from 88 patients).

Two predefined evaluations were performed on the test set:

- Evaluation A compared AI-derived ventricular volume changes with radiology report classifications (decreased/stable/increased), including a threshold-based perceptual sensitivity analysis.
- Evaluation B analyzed patients undergoing ventriculoperitoneal (VP) shunt placement, comparing pre- and postoperative scans. All patients from Evaluation B were also included in Evaluation A.

A detailed study flowchart is provided in Figure 1.

**Figure 1.**
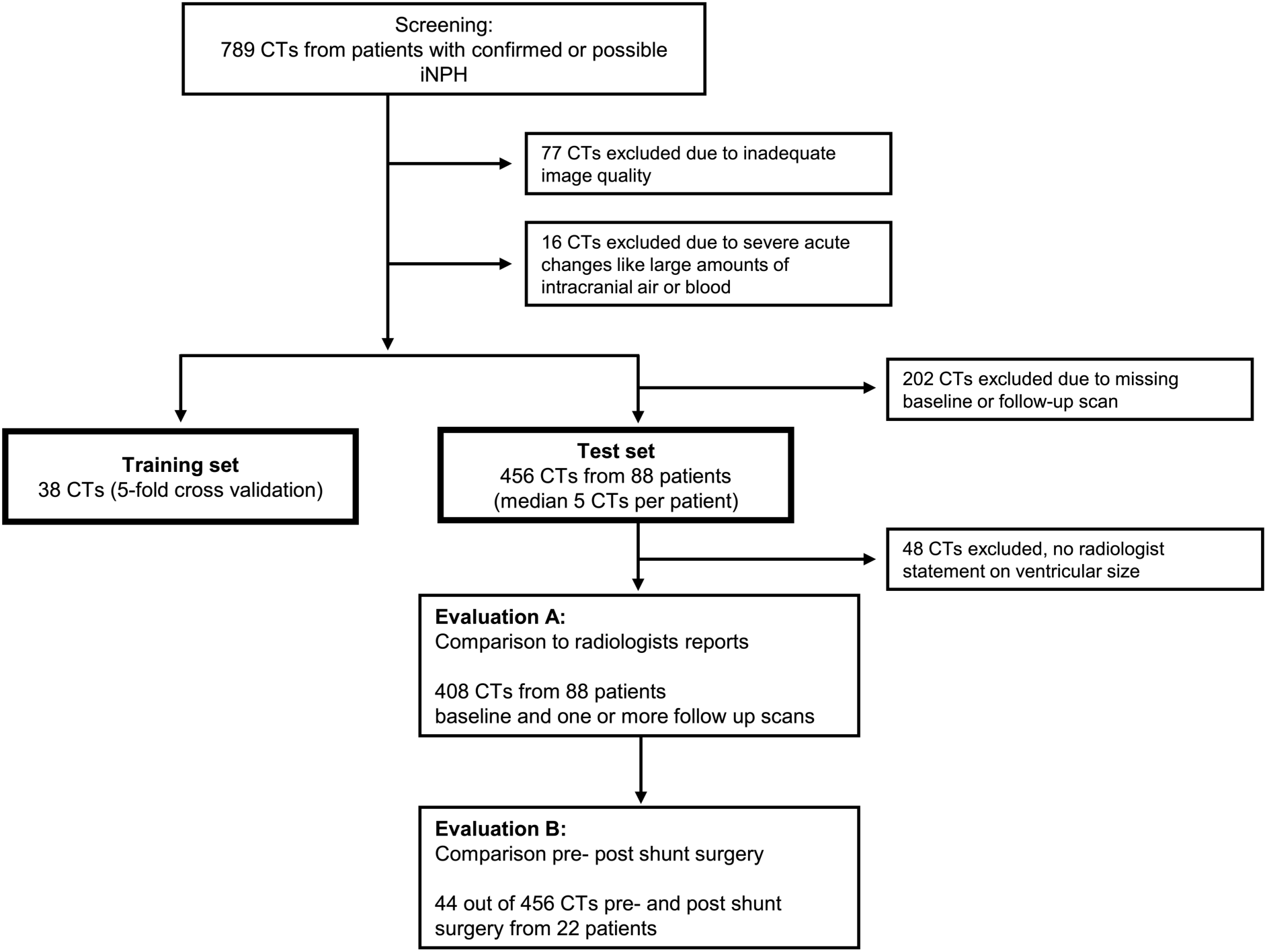
Study flowchart.

Inclusion criteria were:

- Adult patients (aged ≥18 years) diagnosed with probable or possible iNPH according to clinical reports.
- Availability of routine non-contrast-enhanced computed tomography (CT) scans of the head.

Additional inclusion criteria for the test dataset were:

- Availability of a radiology report.
- Each patient had at least two longitudinal CT scans (baseline and follow-up).
- For patients who underwent ventriculoperitoneal (VP) shunt placement, availability of at least one pre- and postoperative CT scan.

Exclusion criteria were:

- Presence of extensive postoperative changes such as large subdural hematomas or extensive intracranial air, obscuring ventricular anatomy.
- Poor-quality CT scans with artifacts.
- CT scans acquired with intravenous contrast enhancement.
- CT scans with slice thickness greater than 1 mm.

### Imaging Acquisition

All CT scans were performed using a Siemens SOMATOM Definition AS 64-slice CT scanner (Siemens Healthineers, Erlangen, Germany). Non-contrast-enhanced axial CT images of the head were acquired with typical parameters (120 kV and 270 mAs) and a slice thickness of ≤1 mm.

### Manual Segmentation and anatomy

Voxel-wise manual segmentation of the ventricular system was performed on the training dataset using the medical image editing software NORA[10]. The ventricles were segmented into 12 anatomical subdivisions: the frontal horns, bodies, occipital horns, temporal horns, and trigones of the lateral ventricles (left and right sides), as well as the third ventricle (including the aqueduct) and the fourth ventricle (Figure 2). Initial segmentation was performed by a medical doctor with one year of experience. A board-certified neuroradiologist, with more than 10 years of experience, reviewed and refined the segmentations to ensure accuracy.

**Figure 2.**
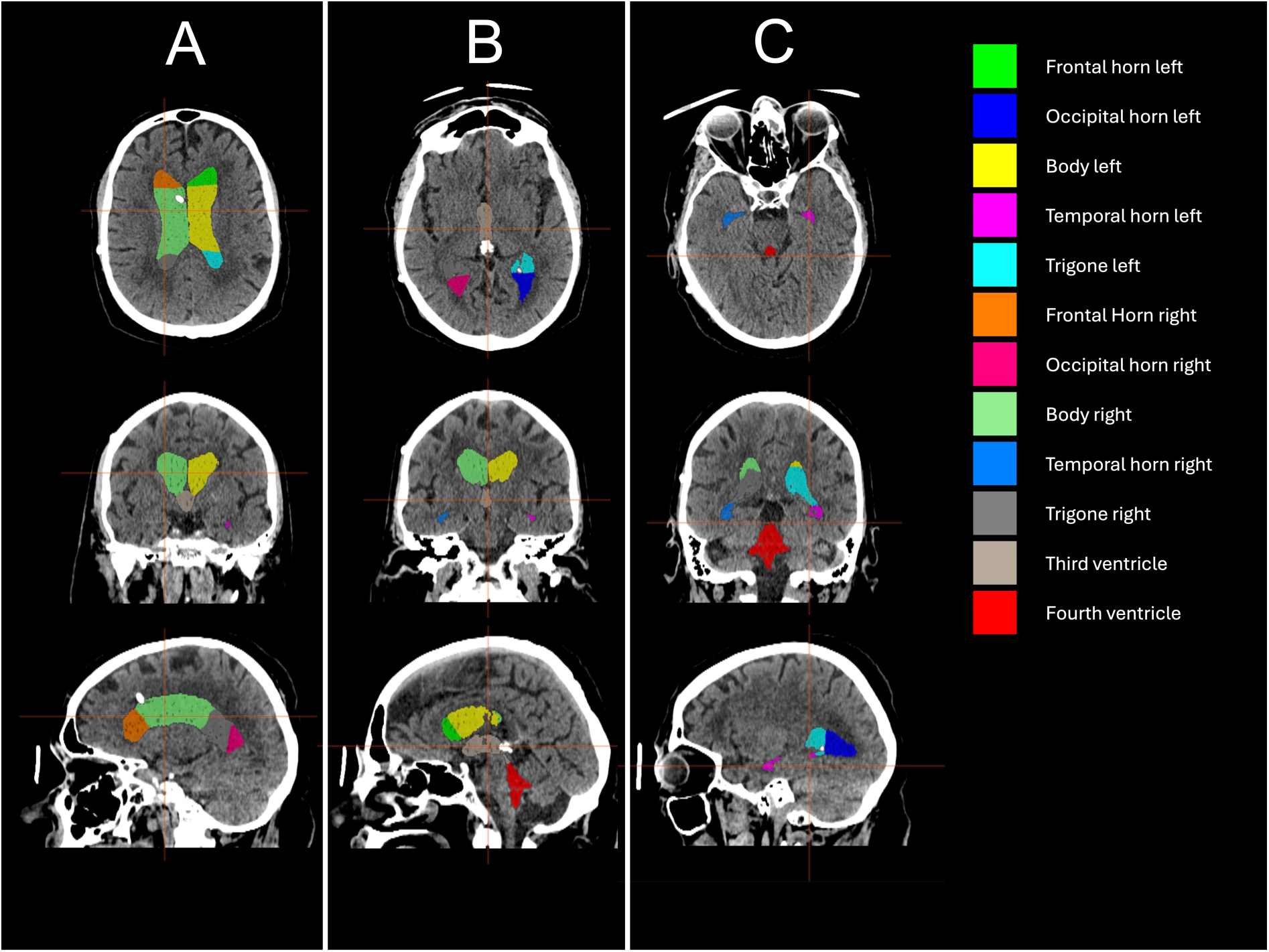
Multi-planar CT views of a representative patient demonstrating excellent automated segmentation of all ventricular subcompartments. Axial, coronal, and sagittal slices are shown for three exemplary, connected levels (A–C), illustrating consistent delineation of all 12 ventricular subcompartments. Note ventricular shunt placed in the right frontal horn, visible in column A (axial and sagittal).

Anatomical landmarks were used based on established neuroanatomical references[11,12]. However, the exact anatomical boundaries of the ventricular subdivisions are not consistently defined in the literature. Notably, the transition between the trigone and the adjacent horns is a gradual and variable boundary. In our segmentation, we used a simple straight superior to inferior line as boundary (see Figure 2).

### Model training using nnU-Net

An AI model based on the nnU-Net framework was used for automated ventricular segmentation. The nnU-Net is a self-configuring deep learning framework for biomedical image segmentation tasks[13].

We utilized a five-fold cross-validation strategy for model training and evaluation. The manually segmented dataset of 38 CT scans was randomly divided into five subsets of approximately equal size. For each fold, the model was trained on 80% (n = 30) of the data and validated on the remaining 20% (n = 8), ensuring that each scan was used for validation exactly once.

Training was performed using the default nnU-Net 3D full-resolution configuration. The framework automatically determined all hyperparameters based on the dataset characteristics. No manual hyperparameter tuning or preprocessing was conducted. Image preprocessing followed the default nnU-Net pipeline (including resampling and intensity normalization). Training was executed on a workstation equipped with a NVIDIA A100 Tensor Core GPU.

### Model Evaluation and volume measurement

The performance of the automated segmentation was assessed using the Dice similarity coefficient (Dice score), calculated between the AI-generated segmentation and the manual ground-truth segmentation for each ventricular compartment. Model performance was evaluated using five-fold cross-validation on the training dataset. Each scan was used for validation exactly once, and Dice scores were averaged across the five folds, resulting in evaluation on 37 training scans (one scan excluded due to segmentation failure).

As the nnU-Net automatically defines all model hyperparameters, no explicit hyperparameter optimization was performed, and no additional hold-out validation set beyond cross-validation was required. For the final application to the testing dataset, the five models trained in each cross-validation fold were ensembled by averaging their output probabilities to generate the segmentation used in the analysis.

Automated ventricular volumes were derived from AI segmentations for all CT scans in the test dataset. Subcompartment volumes in millilitres were calculated from segmentation masks and voxel dimensions, and longitudinal changes were computed between chronologically consecutive scans for each patient.

### Qualitative segmentation evaluation

A fellowship-trained senior neuroradiologist (10 years of experience) evaluated a subset of n=100 AI segmented cases from the test dataset to assess segmentation quality with a focus on clinical applicability.

The subset included 20 cases each where the model detected the largest increase and the largest decrease to the previous scan but deviating from the radiology report stating no change. Additionally, 20 cases each with the largest overall detected decrease, increase, and 20 randomly selected baseline cases were rated.

Each case was rated on a 5-point Likert scale: 5 - Excellent: Perfect segmentation; indistinguishable from manual segmentation by an expert. 4 - Good: Minor deviations, but clinically acceptable with no significant impact on interpretation. 3 - Acceptable: Noticeable errors, but overall segmentation is still usable for clinical purposes. 2 - Poor: Significant errors that affect clinical usability. 1 - Unusable: Segmentation is grossly incorrect and not clinically usable. The neuroradiologist was blinded to the AI volume measurements and previous reports.

### AI quantified volume changes and comparison to radiologist report change

The routine report by the radiologist for each scan was classified into three categories based on statements about ventricular size relative to the previous scan: decreased, stable, or increased. Scans without such a statement were excluded.

For each ventricular compartment, the AI-model derived absolute and percentage volume changes were calculated for the consecutive scans. These values were compared across the three radiologist report categories using a Kruskal–Wallis test, followed by pairwise Mann–Whitney U tests with Bonferroni correction to assess whether AI-measured changes corresponded to radiologist assessments. Because multiple scans per patient were included, results may be influenced by within-patient correlation.

### Threshold-based perceptual sensitivity analysis

For this analysis, two consecutive scans of the same patient were compared. For each pair, the relative ventricular volume change was calculated using the AI and defined as the reference standard.

The AI-derived volume changes were grouped into bins of 3% intervals (e.g., >−6% to −3%, >−3% to 0%, >0% to +3%, >+3% to +6% …). For each scan pair, the corresponding radiologist classification (“increased”, “stable”, or “decreased”) was recorded.

For positive volume changes (>0%), a radiologist classification of “increased” was a correct detection, while “stable” or “decreased” were considered missed detections. For negative volume changes (<0%), a classification of “decreased” was considered a correct detection, while all other classifications were considered missed detections.

Within each bin, sensitivity was calculated as the proportion of correct detections from radiologists among all cases in that bin.

For example, a volume change of −4% falls into the −6% to −3% bin. If the radiologist reported “decreased”, this was counted as a correct detection. If “stable” or “increased” was reported, it was counted as a missed detection.

Finally, we determined the minimum AI-measured volume change at which radiologists identified a correct and corresponding change in at least 50% and 90%. These values were defined as the perceptual detection thresholds.

### Subcompartment Analysis after shunt placement

To evaluate whether percentage volume changes differed systematically across 12 ventricular subcompartments, as well as the right and left lateral ventricle and the total ventricular volume following ventriculoperitoneal (VP) shunt placement, we performed an analysis of covariance (ANCOVA) with an F-test. The dependent variable was the percentage change in volume per subcompartment. The primary fixed effect was subcompartment, with age and sex included as covariates to control for potential confounding.

In case of a significant main effect (p ≤ 0.05), pairwise post-hoc comparisons were conducted using the Tukey Honest Significant Difference (HSD) test.

To visualize adjusted group-level effects while controlling for age and sex, estimated marginal means (EMMs) were computed from the ANCOVA model. These represent the predicted mean percentage volume change per subcompartment, standardized to the overall sample mean of age and sex.

All analyses were performed using Python (v3.8), with statsmodels, pingouin, and scikit-learn libraries. Data distribution was assessed via the Shapiro–Wilk test. Depending on normality, results are presented as either mean ± standard deviation or median (IQR). Statistical significance was defined as p ≤ 0.05.

## Results

### Study Population

A total of 789 non–contrast-enhanced head CT scans from 92 patients met the inclusion criteria for iNPH (Figure 1). 77 scans were excluded because of poor image quality or artifacts and 16 because of severe acute changes obscuring ventricular anatomy, such as excessive intracranial air or hemorrhage. Of the remaining 696 CT scans, 38 were used for model training. An additional 202 scans were excluded because of missing baseline or follow-up examinations.

The final test dataset comprised 456 CT scans from 88 patients (mean age 83.6 years; range 49–102 years; 38 women).

Among these patients, 22 underwent ventriculoperitoneal shunt placement and had both pre-and postoperative CT scans available (median age 82.5 years; IQR 77–87 years). Postoperative imaging was performed between 3 days and 31 months after surgery (median 6 days). All shunts were placed via a right frontal burr hole.

### Performance of the AI Segmentation Model

The AI model based on the nnU-Net framework successfully segmented the ventricles in 37 CT scans with one processing failure in the training dataset. The mean Dice similarity coefficient across all ventricular structures was 0.829, indicating high segmentation accuracy.

The model achieved high segmentation accuracy across ventricular substructures with high mean Dice scores for the fourth ventricle (0.903) and the third ventricle (0.887). The lateral ventricle bodies achieved mean Dice scores of 0.889 (left) and 0.864 (right), the frontal horns 0.877 (left) and 0.859 (right), the occipital horns 0.809 (left) and 0.802 (right), the temporal horns 0.790 (left) and 0.801 (right), and the trigones 0.829 (left) and 0.762 (right).

Representative examples of ventricular segmentation are shown in Figure 2. While concordance with ground truth was high, occasional undersegmentation and inclusion of adjacent hyperdense material (e.g., intraventricular blood) remained challenging (Figure 3).

**Figure 3.**
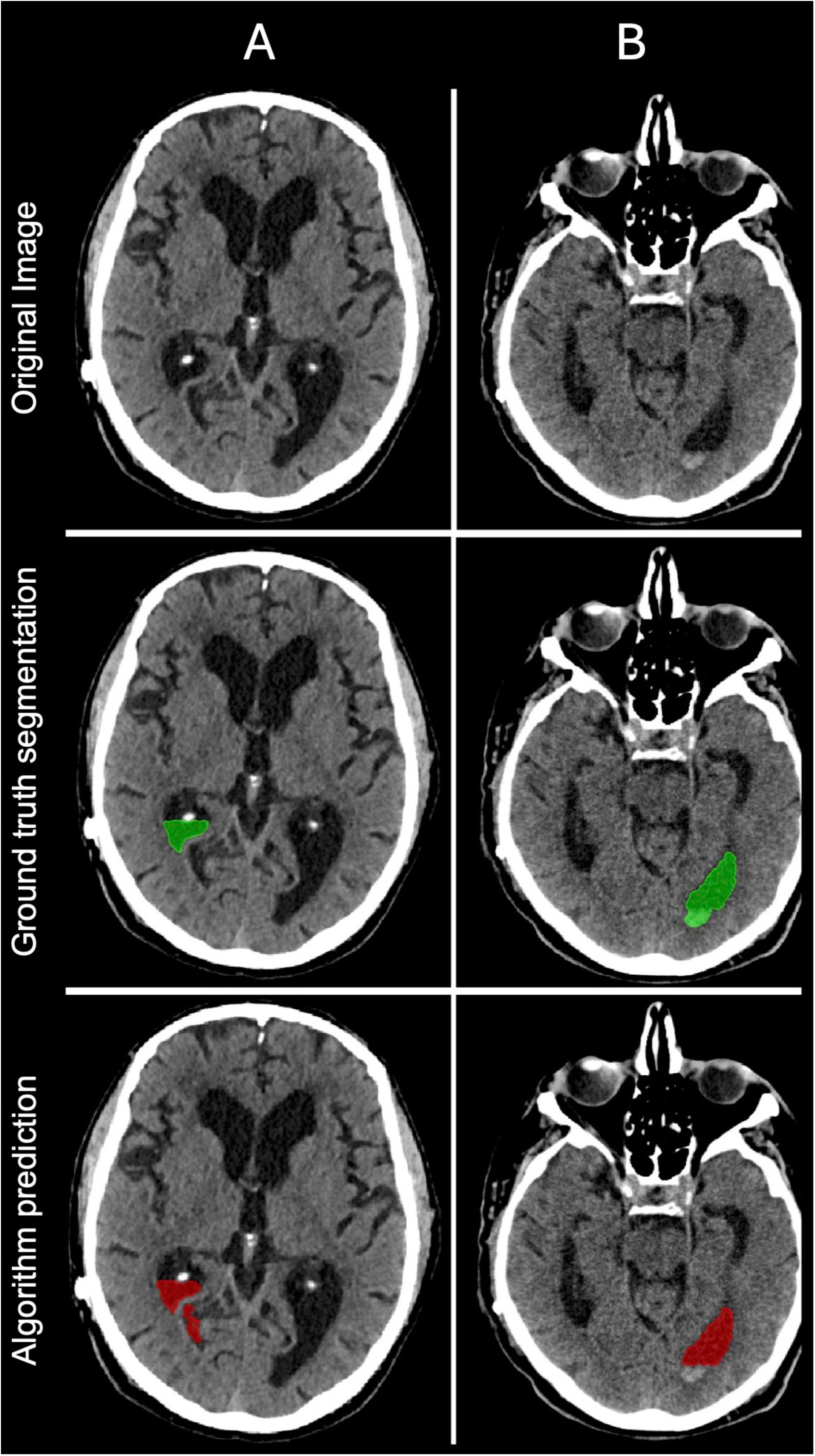
Prediction errors in two different patients (A and B). (A) Partial mis-segmentation of the right occipital horn, with inclusion of parts of the adjacent subarachnoid space. (B) Exclusion of a small amount of intraventricular blood located within the left occipital horn. Although it could be debated whether intraventricular blood should be counted toward the ventricular volume, the model consistently segments only the fluid-filled ventricular space.

### Qualitative Segmentation Evaluation

In total, 100 AI-segmented CT scans were qualitatively assessed by a neuroradiologist. The overall segmentation quality was rated very high, with a median score of 5 (IQR 5–5). Of these, 91 cases (91%) were rated as “excellent” (score 5), 6 cases (6%) as “good” (score 4), and 3 cases (3%) as “acceptable” (score 3).

In the subgroup of 20 randomly selected baseline examinations, the median score was also 5, with 17 cases rated “excellent,” 2 “good,” and 1 “acceptable.”

For the 20 cases with the largest volume increase compared to the previous scan, segmentation quality remained high: 19 were rated “excellent” and 1 “good.” Similarly, in the 20 cases with the largest volume decrease, 19 were rated “excellent” and 1 “good.”

In the subset of scans where the AI detected a change in ventricular volume that was not reported by the radiologist, segmentation quality was again robust. Among the 20 cases with the largest increases, 19 were rated “excellent” and 1 “acceptable.” For the 20 largest decreases, 18 were rated “excellent,” 1 “good,” and 1 “acceptable.”

### Comparison with Radiologist Assessments

Routine radiologic reports of all CT scans in the test dataset were reviewed. Ventricular size was qualitatively described by radiologists as decreased in 68 cases (15 %), unchanged in 287 cases (63 %), increased in 53 cases (12 %), and was not mentioned in 48 cases (10 %).

AI-derived ventricular volume changes showed an excellent correspondence with the qualitative ratings in routine radiology reports (Figure 4). Median relative total ventricular volume change was –17 % (IQR –23 to –10 %; –19 mL, IQR –36 to –11 mL) in cases reported as decreased, 0 % (IQR –5 to 3 %; 0 mL, IQR –6 to 4 mL) in unchanged, and +22 % (IQR 10 to 45 %; +17 mL, IQR 11 to 29 mL) in increased cases.

**Figure 4.**
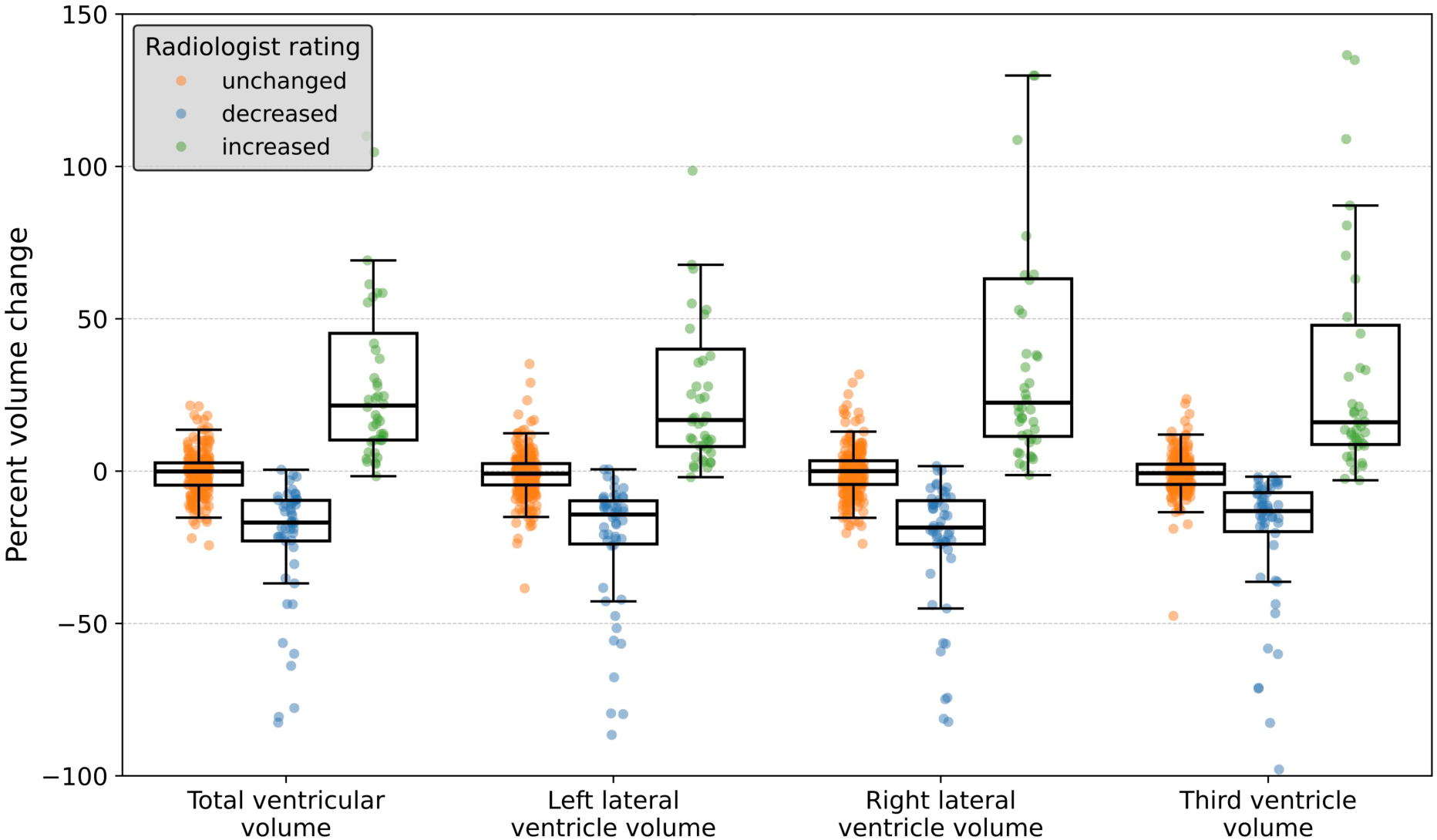
AI-quantified percent volume changes across radiologist-reported categories. Boxplots show AI-measured percent ventricular volume change for CT scans reported by radiologists as unchanged, decreased, or increased. Each dot represents one follow-up scan.

Relative changes were most pronounced in the temporal and occipital horns. In decreased cases, median reductions reached −30% (left temporal horn) and −22% (right occipital horn). In increased cases, the temporal, occipital, and frontal horns showed the greatest enlargement, with increases up to 38%. Changes in the third and fourth ventricles were minor and variable. Across all ventricular subcompartments, the Kruskal–Wallis test confirmed significant differences between the three radiologist categories (p < 0.001), with post-hoc pairwise comparisons revealing consistently robust distinctions between all groups (all corrected p < 0.001).

Full results are shown in the Supplement.

### Perceptual Sensitivity Analysis

A sensitivity analysis was conducted to assess the relationship between AI-detected relative total ventricular volume changes and radiologists’ classifications of enlargement or reduction (Figure 5). Radiologists classified 50% of cases as “increased” when AI detected a volume increase of at least 6.0% (sensitivity: 0.57). At a threshold of 21.0% volume increase, over 90% of cases were classified as “increased” (sensitivity: 0.92). Similarly, 50% of cases were classified as “decreased” when the AI-detected volume decreased by at least 6.0% (sensitivity: 0.52). A decrease of 18.0% or more corresponded to over 90% of cases being classified as “decreased” (sensitivity: 0.92).

**Figure 5.**
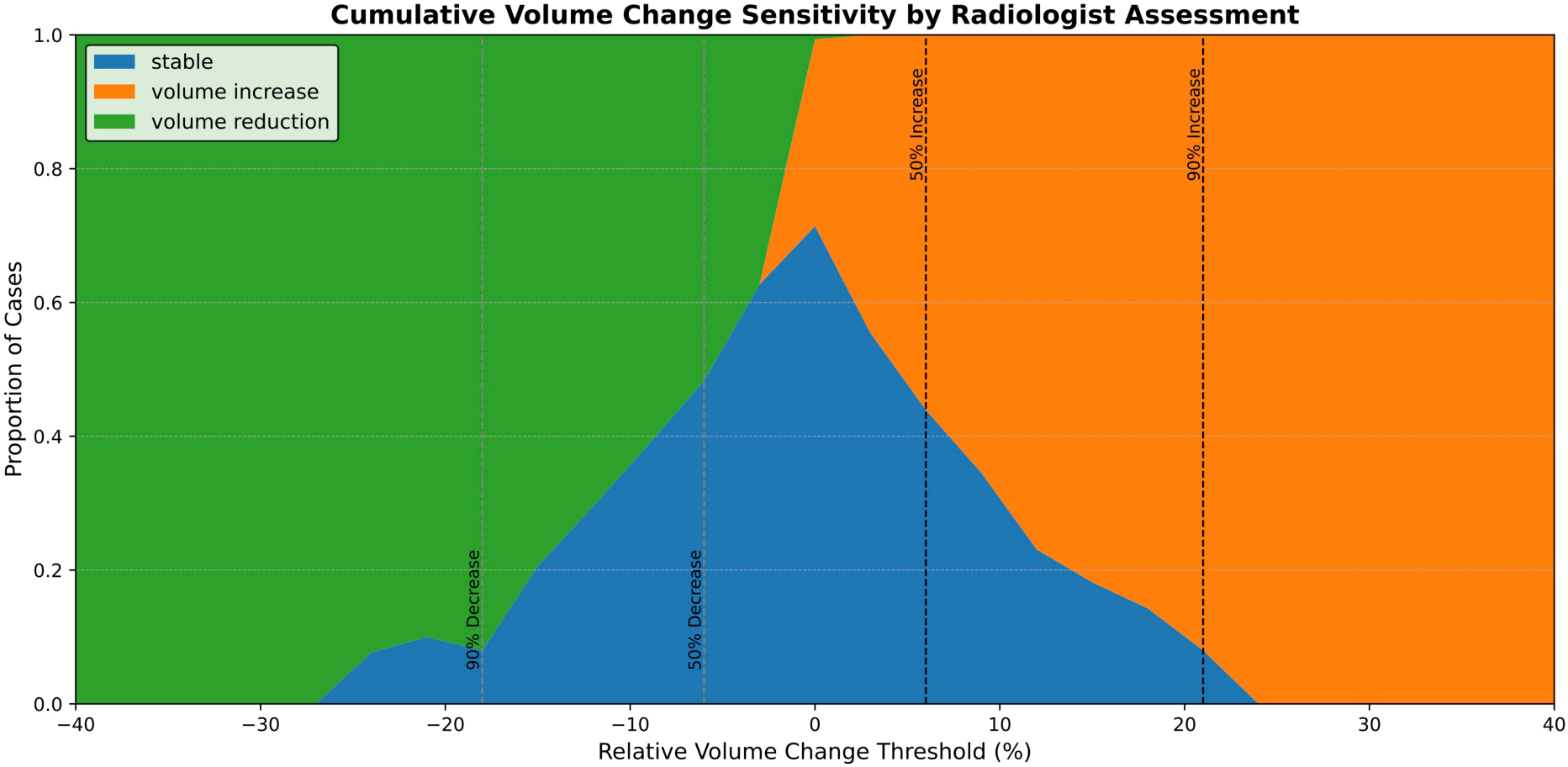
Cumulative volume change sensitivity by radiologist assessment. The x-axis shows increasing relative ventricular volume change as quantified by the AI segmentation model. Colored areas represent the proportion of cases classified by radiologists as decreased (green), unchanged (blue), or increased (orange). Vertical dashed lines indicate the AI-derived volume change thresholds at which radiologists reported change in 50% and 90% of cases.

### Ventricular Volume Measurements Before and After Shunt Placement

The largest relative volume reductions after shunt placement were observed in the right temporal horn (−14%, IQR −25% to −7%) and the right occipital horn (−14%, IQR −16% to −5%), whereas the smallest changes occurred in the left frontal horn (−3%, IQR −7% to +3%) and the fourth ventricle (−2%, IQR −7% to +6%). Overall, total ventricular volume decreased by a median of 8% (IQR −11% to −4%), with greater reductions in the right compared with the left lateral ventricle (−10% vs. −7%).

The ANCOVA revealed a significant main effect of ventricular subcompartment on percentage volume change (F(14, 313) = 3.14, p < 0.01), indicating regional heterogeneity of postoperative volume reduction. Age was a significant covariate (p = 0.03), whereas sex was not (p = 0.1). Post-hoc analysis showed significantly greater reduction in the right temporal horn compared with the fourth ventricle, left frontal horn, and left ventricular body, as well as greater reduction in the right occipital horn and trigone compared with the fourth ventricle (all p ≤ 0.04). No other subcompartmental differences reached statistical significance.

A detailed overview of subcompartmental volume changes is provided in the Supplement and Figure 6.

**Figure 6.**
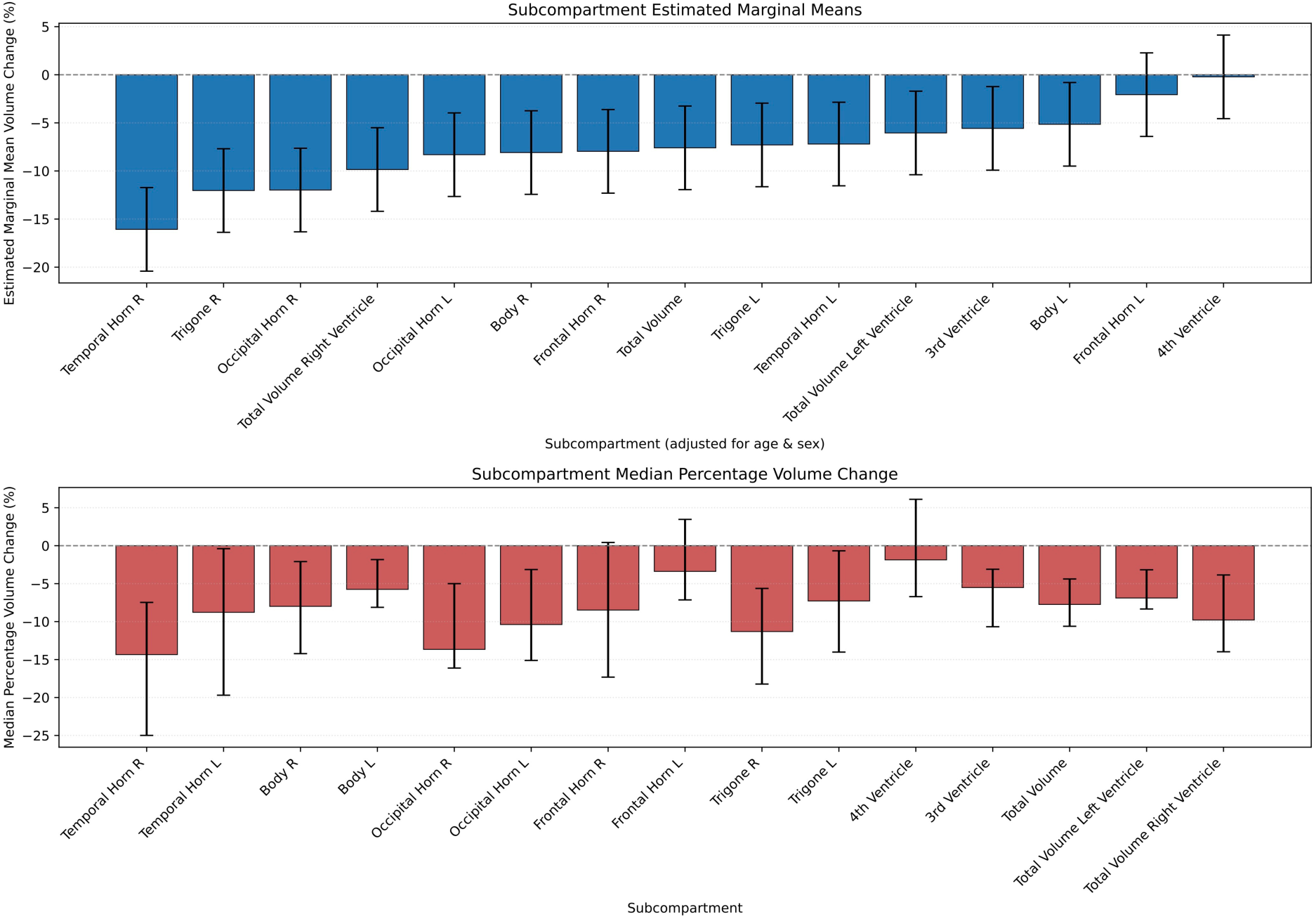
Percentage ventricular volume changes after shunt placement including subcompartments. Estimated marginal means ordered by magnitude (upper panel), adjusted for age and sex. Median percentage ventricular volume changes (lower panel).

## Discussion

This study demonstrates the feasibility and high accuracy of a fully automated, deep-learning nnU-Net[13] based segmentation model for the cerebral ventricular system, including subcompartments, on CT scans in patients with iNPH. With a mean Dice score of 0.83 and a 91% excellent rating on qualitative human radiologist review, the model consistently detects volumetric changes on longitudinal and follow-up imaging, including pre- and post-shunt surgery. The model is available open-source, facilitating the translation from research to real-world clinical implementation.

A recent meta- and systematic review[6] highlights several CT-based studies on AI-based automated ventricular segmentation in hydrocephalus patients, though not exclusively in iNPH[14–22]. These studies primarily provided total ventricular volume measurements with very limited subcompartment delineation, distinguishing only between the lateral, third, and fourth ventricles in some cases[17,20]. Moreover, none of these studies released fully open-source models, with only partial data access available[16,19]. When reported, Dice scores ranged from 0.81[21] to 0.95[14], with most falling above 0.9. While our mean Dice score of 0.83 across all subcompartments is slightly lower, this likely reflects a broader subcompartment variability, where precise segmentation is more challenging, such as in the boundaries of the trigone. Notably, none of these studies evaluated the models against human qualitative judgment. Furthermore, our model was assessed on one of the largest reported iNPH cohort in CT AI studies, comprising 88 patients.

We evaluated the model in two real-life, clinical scenarios:

First, automated volumetry showed strong correspondence with radiological assessments, consistently detecting increased, decreased, or unchanged ventricular volume on follow-up imaging as reported by radiologists. The model detected subtle changes (about ±6% of total ventricular volume), yet radiologists labeled only about half of these cases as changed, while changes with an increase above 21% and a decrease below 18% compared to the previous scan were reported as changed in >90% of cases by the radiologist. This suggests a perceptual threshold in routine readings.

Prior iNPH studies show that visual assessment frequently underestimates or misclassifies volumetric changes compared with automated measurements[5]. These discrepancies likely reflect the limitation of visual change detection in complex 3D anatomy, where subcompartments change at different rates (most prominently the temporal, occipital, and frontal horns), as well as inherent variability in CT interpretation. Although the AI segmentation demonstrated excellent performance, treating it as a gold standard may overlook potential segmentation errors. Automated volumetry should therefore be considered a complementary tool to stabilize longitudinal assessment and indicate trends rather than replace radiological judgment.

Second, after ventriculoperitoneal shunt placement, the model identified a mean total ventricular volume reduction of −8%. The largest changes involved the temporal and occipital horns, with greater reductions on the right compared with corresponding left-sided compartments, consistent with right-sided shunt placement.

Subcompartment-specific changes are rarely reported, but disproportionate temporal horn enlargement has been described[23]. Quantitative ventricular volumetry has been reported to correlate more closely with postoperative clinical improvement than linear indices such as the Evans Index[24], suggesting that higher subcompartmental resolution may identify novel volumetric patterns associated with shunt responsiveness and outcome.

These two scenarios demonstrate the real-world clinical applicability of our model, offering a consistent tool for follow-up and longitudinal evaluation of ventricular volume, potentially reducing variability in radiologist-dependent assessments.

Limitations include the retrospective single-center design without external validation, exclusion of complex postoperative cases, and limited sample size for pre-/post-shunt analysis. Also, radiological change assessment is perceptual, and small discrepancies are expected. Multicenter validation, clinical outcome correlation, and workflow integration remain to be evaluated.

## Conclusion

This study shows that fully automated AI-based ventricular segmentation on head CT has high accuracy and clinical consistency in patients with iNPH. Automated volumetry provides reproducible quantification of ventricular change, matching with radiological assessment and offering greater sensitivity to subtle volume changes. Reliable detection of postoperative subcompartmental changes supports its role as an adjunct for longitudinal monitoring and potentially therapeutic decision-making.

## Declarations

### Ethics approval

This retrospective study was conducted in accordance with the Declaration of Helsinki. Ethical approval was waived by the local ethics committee due to the retrospective design and use of fully anonymized data.

### Consent to participate

Informed consent was waived by the local ethics committee due to the retrospective design and use of fully anonymized data.

### Consent to publish

Not applicable.

### Code availability

The model is available as open-source software at https://github.com/wasserth/TotalSegmentator and https://totalsegmentator.com/

### Author contributions

All authors contributed to the study conception and design. Material preparation, data collection and analysis were performed by MAM, SAG, JW, AKI, NV, MB, JML. The first draft of the manuscript was written by MAM, SAG and JML and all authors commented on previous versions of the manuscript. All authors read and approved the final manuscript.

## Data Availability

All data produced in the present study are available upon reasonable request to the authors

The datasets generated and/or analyzed during the current study are not publicly available due to patient privacy and institutional restrictions but may be available from the corresponding author upon reasonable request, subject to institutional approval.

